# Randomized Controlled Comparative Effectiveness Trial of Risk Model-Guided Clinical Decision Support for Suicide Screening

**DOI:** 10.1101/2024.03.14.24304318

**Authors:** Colin G. Walsh, Michael A. Ripperger, Laurie Novak, Carrie Reale, Shilo Anders, Ashley Spann, Jhansi Kolli, Katelyn Robinson, Qingxia Chen, David Isaacs, Lealani Mae Y. Acosta, Fenna Phibbs, Elliot Fielstein, Drew Wilimitis, Katherine Musacchio Schafer, Rachel Hilton, Dan Albert, Jill Shelton, Jessica Stroh, William W. Stead, Kevin B. Johnson

## Abstract

Suicide prevention requires risk identification, appropriate intervention, and follow-up. Traditional risk identification relies on patient self-reporting, support network reporting, or face-to-face screening with validated instruments or history and physical exam. In the last decade, statistical risk models have been studied and more recently deployed to augment clinical judgment. Models have generally been found to be low precision or problematic at scale due to low incidence. Few have been tested in clinical practice, and none have been tested in clinical trials to our knowledge.

**Methods:** We report the results of a pragmatic randomized controlled trial (RCT) in three outpatient adult Neurology clinic settings. This two-arm trial compared the effectiveness of Interruptive and Non-Interruptive Clinical Decision Support (CDS) to prompt further screening of suicidal ideation for those predicted to be high risk using a real-time, validated statistical risk model of suicide attempt risk, with the decision to screen as the primary end point. Secondary outcomes included rates of suicidal ideation and attempts in both arms. Manual chart review of every trial encounter was used to determine if suicide risk assessment was subsequently documented.

**Results:** From August 16, 2022, through February 16, 2023, our study randomized 596 patient encounters across 561 patients for providers to receive either Interruptive or Non-Interruptive CDS in a 1:1 ratio. Adjusting for provider cluster effects, Interruptive CDS led to significantly higher numbers of decisions to screen (42%=121/289 encounters) compared to Non-Interruptive CDS (4%=12/307) (odds ratio=17.7, p-value <0.001). Secondarily, no documented episodes of suicidal ideation or attempts occurred in either arm. While the proportion of documented assessments among those noting the decision to screen was higher for providers in the Non-Interruptive arm (92%=11/12) than in the Interruptive arm (52%=63/121), the interruptive CDS was associated with more frequent documentation of suicide risk assessment (63/289 encounters compared to 11/307, p-value<0.001).

**Conclusions:** In this pragmatic RCT of real-time predictive CDS to guide suicide risk assessment, Interruptive CDS led to higher numbers of decisions to screen and documented suicide risk assessments. Well-powered large-scale trials randomizing this type of CDS compared to standard of care are indicated to measure effectiveness in reducing suicidal self-harm.

ClinicalTrials.gov Identifier: NCT05312437

## Introduction

Improving suicide prevention requires appropriate risk identification, prognostication, and effective intervention. Risk identification combines clinical judgment, validated screening instruments, and a growing cadre of validated statistical models.^1,2^ Computational risk prediction might be suited to prompt further suicide risk assessment and/or intervention, but effectiveness of model-driven clinical decision support (CDS) systems in suicide prevention is understudied.^2–4^ The most prominent pre/post evaluation of a preventive outreach program, REACH VET, remains an exemplar to date, though no such system has been studied via randomized controlled trial (RCT) to our knowledge.^5^

Traditional suicide risk prognostication relies on clinical judgment guided by validated instruments like the Patient Health Questionnaire,^6^ Columbia Suicide Severity Rating Scale (CSSRS),^7^ the Ask Suicide-Screening Questions toolkit,^8^ and others.^9,10^ In the last decade, a myriad of validated statistical models have been published to improve suicide prognostication.^1^ These include Army STARRS,^11–13^ REACH VET,^5,14,15^ the Mental Health Research Network,^16,17^ and many more.^18–23^ Recent research suggests that statistical modeling combined with face-to-face screening outperform either alone.^24^

To enable prevention, predictive models must be actualized through tools like CDS. Prior literature outside suicide research has examined forms of CDS such as interruptive (e.g., alerts) and non-interruptive (e.g., static icons or visual cues) to inform contact isolation decisions,^25^ laboratory alerts,^26^ and blood transfusion.^27^ While interruptive CDS tends to be more effective in driving behavior, this question has not been studied via RCT in suicide preventive workflows to our knowledge. Also, given significant concerns around false positives in suicide screening,^28,29^ demonstrating adequate performance of a non-interruptive CDS would support implementing a less burdensome and stigmatizing alert.^30,31^

Our team has previously validated, replicated, and prospectively “silently” tested an Electronic Health Record (EHR)-based suicide risk model.^24,32,33^ Here, we report design and evaluation of CDS driven by that model to prompt suicide risk assessment within healthcare encounters in settings that do not conduct universal screening. The designs of the CDS and the research protocol are informed by Human-Centered Design (HCD),^34^ a framework to evaluate appropriate CDS alerts and responses,^35^ and a deployment framework for clinical artificial intelligence.^36^ We conduct a Comparative Effectiveness RCT (ClinicalTrials.gov Identifier: NCT05312437) of our risk model-prompted CDS, assessing two CDS designs, Interruptive and Non-Interruptive,^25,37,38^ to prompt suicide risk assessment within clinical encounters. We hypothesize the Interruptive CDS arm would lead to higher rates of in-person suicide risk assessment compared to the Non-interruptive CDS arm.

## Methods

This pragmatic, two-arm RCT uses a validated risk model to prompt suicide preventive CDS at the start of routine healthcare encounters.^24,32,33,39^

### Study Setting

A non-behavioral health setting with increased suicide risk^40^ and variable suicide prevention workflows, ambulatory Neurology clinics serve as the trial setting. Unlike high-risk settings such as the Emergency Department, ambulatory Neurology clinics do not have universal screening protocols in all sites. Despite the absence of these protocols, some patients in these settings have increased suicide risk, e.g., those with movement disorders and inherited disorders like Huntington’s Disease.^41,42^

The study settings in this RCT include ambulatory Neurology clinics across three divisions: Neuro-Movement Disorders; Neuromuscular Disorders; Behavioral and Cognitive Neurology. The study was conducted at Vanderbilt University Medical Center, an academic medical center in the Mid-South of the United States.

### Intervention (CDS) Design

We co-designed the CDS with neurologists through multiple meetings with volunteer physicians in the study clinics. Our HCD experts (S.A., C.R., L.N.) with our EHR-physician builder (A.S.) designed two versions of the CDS – Interruptive and Non-Interruptive. In the Interruptive CDS, an alert window (Best Practice Alert or “BPA”) and a patient panel icon are visible simultaneously. “Dismissing” the alert hides it with no effect on the patient panel icon. An eSupplement includes all CDS visualizations.

The Non-Interruptive CDS uses a summarization panel for patient-level data foundational to the EHR interface (“Epic Storyboard”). This design resulted directly from stakeholder input. They asked for an ability to engage with the CDS in an accessible location when a physician notes the CDS but might not be ready to act on it. When relevant, the Non-Interruptive CDS displays, “Elevated Suicide Risk Score” in the patient summarization panel (shown in eSupplement). Hovering over this icon results in a pop-out identical to the Interruptive CDS. Clicking that pop-out permits physicians to act on the alert identically to interaction in the Interruptive CDS arm.

During HCD, physicians requested a means to better document suicide screening assessments within encounters. In response, we developed a customized form (shown in eSupplement) using the CSSRS, our medical center’s chosen instrument for universal screening mandated by the Joint Commission.^43^

Overall CDS logic links interactions directly to trial outcomes (Figure 1).

**Figure 1:**
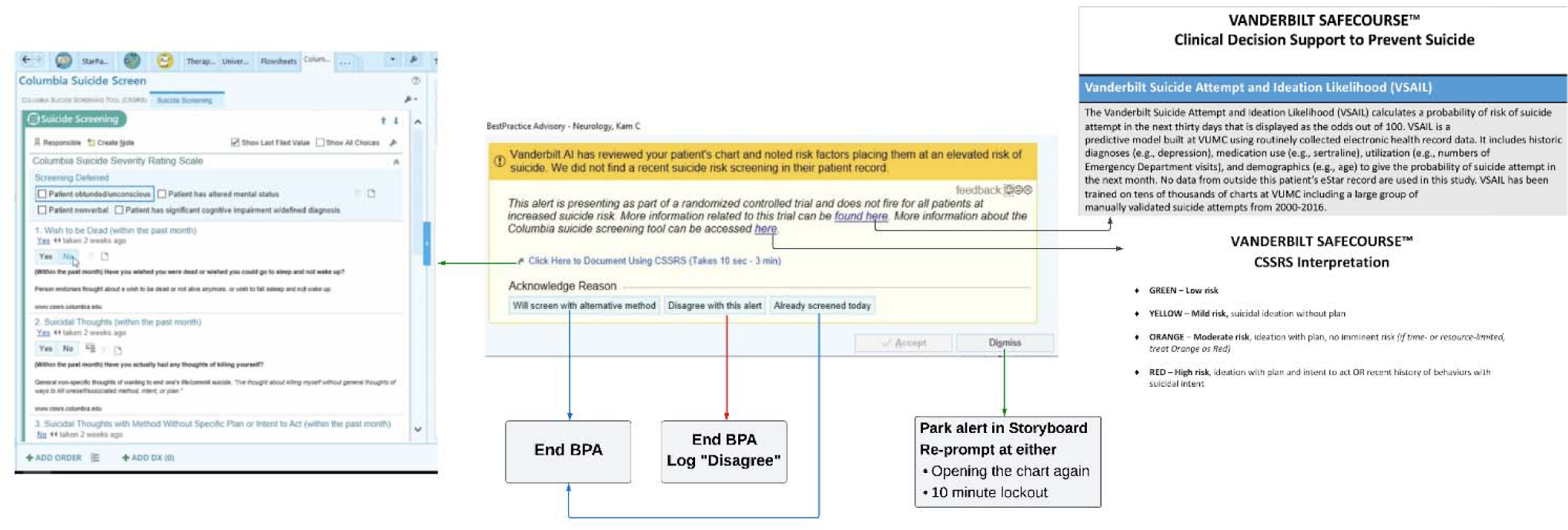
CDS Logic, detailed views of each interface in eSupplement.

### Randomization and masking

During patient check-in/registration for an encounter in-clinic or over telehealth, our validated risk model calculates thirty-day suicide attempt risk (probability) using operational data across diagnoses, medications, visit utilization, and demographics.^32,33^ These probabilities are transmitted to flowsheets in the EHR to prompt CDS. 1:1 randomization occurs for all patients with predicted risk above or equal to 2%, a threshold chosen from prior validation.^33^ Randomization is conducted directly within the EHR with half randomized to Interruptive CDS and the remainder to Non-Interruptive.

The intervention itself reflects randomization status – i.e., interruptive or not, making masking/blinding the intervention infeasible.

### Trial Eligibility

As this trial was pragmatic in design, all patients appearing for routine care in study settings were eligible.

### Primary Trial Outcome and Sample Size

The decision to assess suicide risk in-person through CDS interaction serves as the primary trial outcome. The primary outcome was recorded via direct interaction with study CDS (see Figure 1).

With ∼15 patients per week per arm estimated from silent validation,^33^ we hypothesized the Interruptive CDS would be more effective at prompting in-person suicide risk assessment than Non-Interruptive (20% compared to 5%). Thus, we needed at least 75 patients in each arm to achieve 90% power with 5% probability of type I error.

### Secondary Outcomes

Secondary trial outcomes include rates of thirty-day episodes of suicidal ideation, suicide attempt, rates of documented suicide risk assessment in clinical notes, psychiatric hospitalization, or emergency department utilization related to mental illness and/or suicide risk. We ascertain suicidal ideation and attempts with any documented diagnostic codes or chart review. Diagnostic codes have been shown to have high positive predictive value (PPV) in International Classification of Diseases, version 10, in our prior research including PPV of 0.85 for suicide attempt and 0.96 for suicidal ideation.^44^ We ascertain hospitalization or emergency department utilization with EHR healthcare encounter data.

We assess documentation via chart review of every trial encounter by two members of the study team (J.K., K.R.) with adjudication when needed by the Principal Investigator (C.W.). During the chart review, annotators recorded presence/absence of documented suicide risk assessments and justifying clinical text (“seed terms”, e.g., “Denied SI”).

### Pre/Post Comparator Analysis

Because this RCT compares CDS effectiveness, a standard of care comparator for similar predicted risk patients was assessed from August 2021-February 2022, one year earlier than the RCT. We conducted an identical analysis of notes during this earlier period using the same seed terms extracted during RCT chart review above (see eSupplement for seed terms).

### Statistical Analyses

Chi squared test statistics support primary hypothesis testing. We analyze baseline study characteristics with descriptive statistics and pooled p-values. Cohen’s kappa measures inter-rater agreement in chart review. We summarize baseline variables in counts and frequencies or mean and standard deviation (sd) for categorical or continuous variables, respectively. Chi-squared tests and two-sample t-tests compare variables between the two arms. The primary study outcome, decision to screen, and one of the secondary outcomes, documentation of suicide risk assessment, are assessed with logistic regression models with the Huber-White method to adjust for potential cluster effects by provider.^45^

### Thematic Analysis

Free text comments entered by providers into our CDS are reviewed manually and separately by three members of the study team (S.A., L.N., C.W.) for emergent themes and patterns.

### Training, Outreach, and Education

To increase engagement, acculturate physicians to the RCT, and begin CDS education, our study team engaged clinical teams first through division seminars for the Department of Neurology. Department-wide and targeted email supplemented this effort to increase awareness of the upcoming trial. Educational materials were prepared and disseminated including an instructional video with demonstrations of both Interruptive and Non-Interruptive CDS. Materials were distributed via email to physicians in each trial site. Additionally, materials were stored in a secured document repository and accessible directly from links built into the CDS itself (eSupplement).

### Protocol Deviations

We monitored the trial protocol throughout the study including monthly check-ins with clinical sites and feedback tools built into the CDS. The approved study protocol is available at clinicaltrials.gov^46^ and in supplementary material here (eSupplement).

### Safety and Adverse Events

The study team reviewed trial progress monthly and shared study personnel contact information broadly with participating sites.

### Inclusion and Ethics Statement

Patients already scheduled for neurologic care were enrolled in this trial. Waiver of consent was requested and approved by the VUMC IRB (#210865) given concern for introducing bias into clinical encounters in which providers might credibly disagree with the CDS and decide not to assess suicide risk, which otherwise might be prompted by the consent process itself.

### Ethics Approval

Ethicists were represented on our study team throughout the study design period. The study team met with the Office of Legal Affairs prior to trial start given the sensitive nature of suicide prevention to avoid unintended liability risks to providers.

### Reporting Summary

We registered the RCT in ClinicalTrials.gov in April 2022 (NCT05312437).^46^ At VUMC, we obtained approval or assent from the VUMC IRB (#210865), legal affairs, Health Information Technology CDS committees, and Medical Device Regulatory Affairs Program Review. This RCT manuscript and attached protocol adhere to both CONSORT^47^ and SPIRIT^48^ guidelines for reporting and the study protocol, respectively. Relevant checklists for CONSORT and SPIRIT are eSupplements.

## Results

### Study Sample

From August 16, 2022, through February 16, 2023, our study randomized 596 of 7,732 total encounters (596/7,732=8%) in RCT settings. The trial ended as scheduled with sufficient sample sizes to assess primary outcome differences per arm. The randomized encounters involved 561 of 6,062 total patients (561/6062=9%). Seventy-one providers participated in the trial receiving either Interruptive or Non-interruptive CDS: twenty-four attendings; twenty-six resident physicians; six fellows; six nurse practitioners; five psychologists; three genetic counselors; one physician assistant. The baseline study characteristics at the first study encounter for patients seen in this period are shown (Table 1).

**Table 1:**
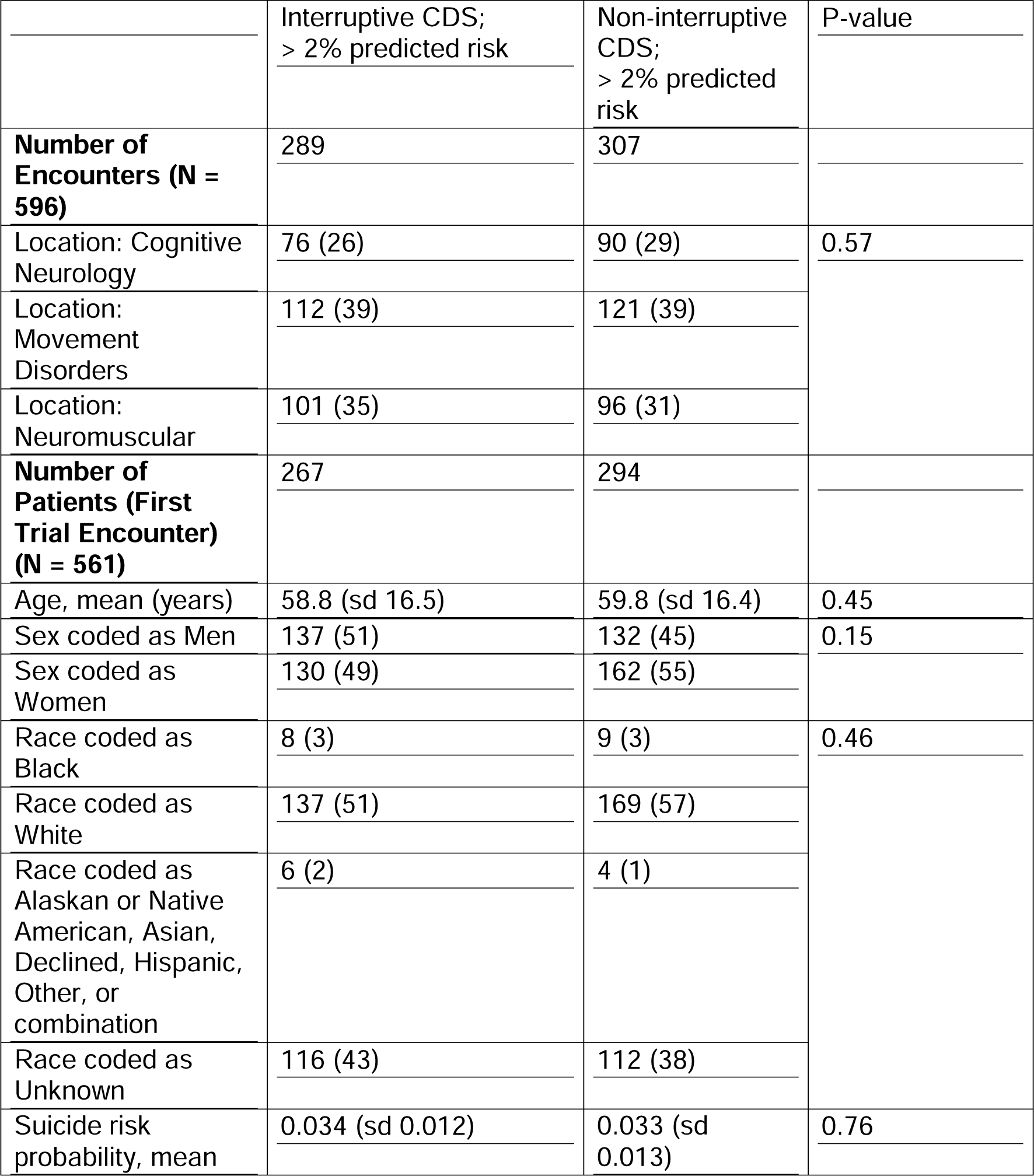
Baseline Study Characteristics, data are N (% unless otherwise noted) Primary trial outcome.

Of 289 encounters in the Interruptive CDS arm, 121 (42%) resulted in providers electing to screen either through the CSSRS or another assessment of providers’ choosing. Of 307 encounters in the Non-interruptive CDS arm, 12 (4%) led to screening with the CSSRS or other assessment. Accounting for cluster effects by individual providers, the Interruptive CDS prompted higher rates of in-person screening compared to non-interruptive CDS (OR=17.7, 95% confidence interval: 6.42-48.79, p-value <0.001), consistent with the study alternative hypothesis. Analyses were conducted by original assigned groups from randomization.

### Secondary trial outcomes

While the proportion of documented risk assessments among those noting the decision to screen was higher for providers in the Non-Interruptive arm (92%=11/12) than in the Interruptive arm (52%=63/121), the Interruptive CDS was associated with more frequent documentation of suicide risk assessment (22%=63/289 encounters compared to 4%=11/307, p-value<0.001). Figure 2 indicates decisions to screen (primary outcome) and documentation rates (secondary) by trial arm.

**Figure 2:**
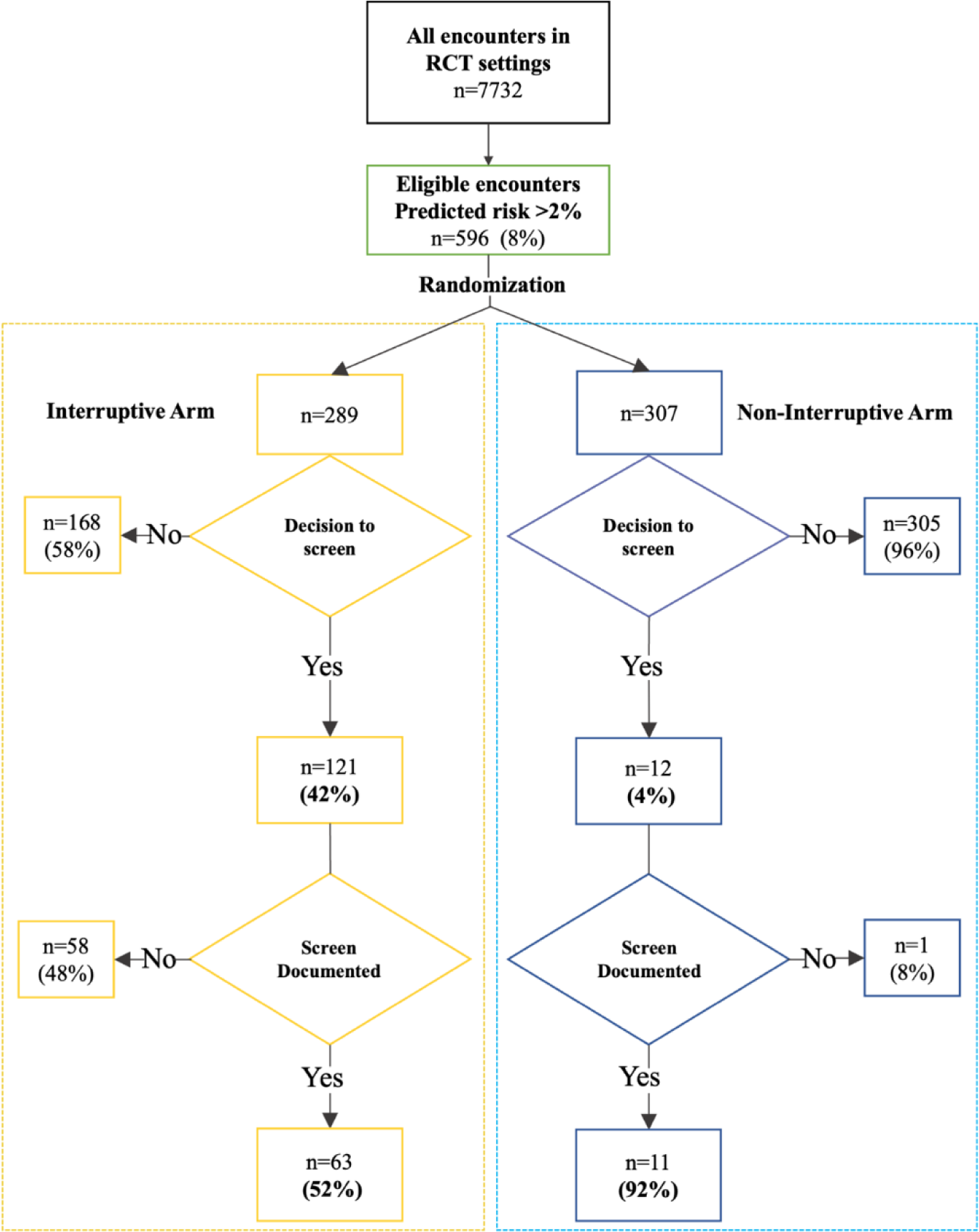
Flowchart of Trial Outcomes by Arm.

The two chart reviewers had excellent agreement in independent, manual chart review to confirm documentation of suicide risk assessment (Cohen’s kappa 0.9, goal > 0.8).

No adverse safety events or protocol deviations were reported throughout the trial.

There were no documented episodes of thirty-day suicidal ideation, suicide attempt, emergency department utilization for mental illness, or psychiatric hospitalizations in either arm.

### Pre-/Post Analysis

From August 2021 through February 2022, the same clinical settings had a baseline suicide risk assessment rate of 7.7% (64 of 832 encounters in the same settings, one year before this RCT).

### Thematic Analysis of Provider Comments

Through the design of the CDS, physicians were able to share comments on use of the alerts in practice. Forty comments were entered out of all study encounters (40/596=6.7%, 13 in the Non-Interruptive and 27 in the Interruptive arm, comments shared in eSupplement). The major themes included whether or not a patient was “Screened” and whether that screen was negative (32 of 40 comments); whether the alert was “Inappropriate for the Patient” (4 of 40 comments); and whether the screening was “Deferred for Patient” (2 of 40 comments).

## Discussion

This pragmatic RCT compared the effectiveness of two versions of CDS prompted by high predicted risk using real-time statistical risk modeling in the clinic. Neurology clinics were chosen as the trial setting because suicide risk is elevated among populations with neurologic disorders, but suicide screening assessment is variably incorporated into neurologic care ^40,49,50^. Interruptive CDS prompted higher absolute numbers of suicide risk assessments. Non-interruptive CDS showed a higher proportion of decisions to screen followed by assessments documented assessment, not simply indicated to the CDS. That is, decisions to screen were more likely to result in a documented assessment in the Non-Interruptive arm though absolute numbers were small. No thirty-day suicide events followed study encounters throughout the trial.

This trial builds on understanding of effectiveness of forms of CDS to drive clinical decisions. Interruptive CDS tends to be more effective in prompting behavior. This finding has been shown in diverse settings including heart failure medication management,^37^ contact isolation practices in the emergency department,^25^ and PHQ9 administration in primary care clinics.^51^ Disadvantages of interruptive CDS, especially alert fatigue, counterbalance its effectiveness.^52,53^ Providers prefer passive, non-interruptive CDS even while acknowledging they might not be seen nor used as often as interruptive prompts.^54^ We note that regardless of the form of CDS, using a validated statistical model to prompt CDS reduced the potential burden of screening to only 8% of all 7,732 encounters in trial settings.

Limitations of this RCT include potential leakage of suicide risk assessment that might have occurred regardless of the presence of CDS. The RCT was not powered to detect changes in rates of suicide attempts or deaths from suicide (neither of which occurred during the RCT). CDS iterative design including integration of treatment recommendations as well as larger-scale RCTs of this type of CDS should be foci of future work.

In this trial, a validated predictive model prompted randomization to two forms of CDS prevention. Interruptive CDS outperformed non-interruptive CDS in prompting in-person assessments. The predictive modeling trigger reflects the need to precisely prompt conversations about suicide risk – a clinical priority that remains rare at the scale of a health-system. Imprecision has been cited as a challenge for both screening instruments^28^ and predictive models in suicide prevention.^29^ While the predictive model here reduced the number of CDS prompts from ∼60 per day (all clinic encounters, were universal screening to be implemented in study settings) to ∼4-6 per day, a larger-scale trial focused on the effectiveness of the overall system powered sufficiently against standard of care to reduce suicide events is indicated. Future research might also further iterate CDS design, test these systems in more diverse settings, and integrate preventive recommendations beyond risk assessment.

## Supporting information

Clinical Decision Support

CONSORT Flow

CONSORT Checklist

Study Protocol

## Data Availability

Because data include sensitive PHI, data are not available for dissemination outside the study team.

## Acknowledgements

This work was supported by the Evelyn Selby Stead Fund for Innovation (Vanderbilt University Medical Center), NIMH R01MH118233, R01MH121455, R01MH116269, R01 MH120122, NHGRI RM1 HG009034, NIH U54 HG012510, FDA Sentinel WO2006, Wellcome Leap MCPsych.

## Data Availability Statement

This study relies on identified, sensitive health records. Our team does not have permission to distribute these data with the manuscript.

## Code Availability Statement

No custom code or mathematical algorithms are central to our conclusions. The analytic code used in this study calculated the results of common statistical tests.

