## Supplementary material for "Randomized Controlled Comparative Effectiveness Trial of Risk Model-Guided Clinical Decision Support for Suicide Screening": CONSORT Flow

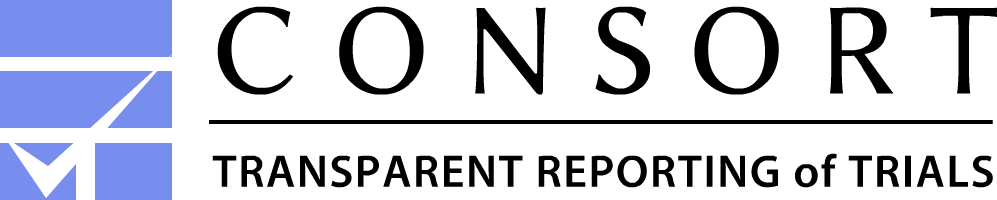

**CONSORT 2010 Flow Diagram**

### Allocation

Allocated to Interruptive CDS (n= 289 )

♦ Will screen with alternative method (N=45, 18 MDs)

♦ Clicked CSSRS Link (N=24, 11 MDs)

♦ Completed CSSRS (N=15, 5 MDs)

♦ Already screened today (N=37, 18 MDs)

♦ Canceled BPA/Ignored (N=107, 16 MDs)

♦ Disagree (N=61, 16 MDs)

Allocated to Non-Interruptive CDS (n= 307 )

♦ Will screen with alternative method (N=6, 2 MDs)

♦ Clicked CSSRS Link (N=0, 0)

♦ Completed CSSRS (N=0, 0)

♦ Already screened today (N=6, 3 MDs)

♦ Canceled BPA/Ignored (N=291, 44 MDs)

♦ Disagree (N=3, 3 MDs)

♦ Accept BPA (no action taken) (N=1, 1 MD)

Randomized (n= 596 encounters)

561 patients

72 providers

Excluded (n=	)

♦  Not meeting inclusion criteria (n= 7,800 )

♦  Declined to participate (n= 0 )

♦  Other reasons (n= 0)

Assessed for eligibility (n= 8,396 )

### Follow-Up

Lost to follow-up (give reasons) (n=0)

Discontinued intervention (give reasons) (n=0)

Analysed (n=307 )
♦ Excluded from analysis (give reasons) (n= 0)

### Analysis

Lost to follow-up (give reasons) (n= 0)

Discontinued intervention (give reasons) (n=0)

Analysed (n= 289 )
♦ Excluded from analysis (give reasons) (n= 0 )

### Enrollment
