## Supplementary material for "Randomized Controlled Comparative Effectiveness Trial of Risk Model-Guided Clinical Decision Support for Suicide Screening": CONSORT Checklist

### Clinical Decision Support to Prevent Suicide:

### Colin G Walsh, MD, MA

2525 W End Ave, Suite 1475

Nashville, TN 37203

Department of Biomedical Informatics

Department of Medicine

Department of Psychiatry and Behavioral Sciences

Vanderbilt University Medical Center

****

**Trial Registration**

ClinicalTrials.gov Identifer: NCT05312437

URL: https://clinicaltrials.gov/study/NCT05312437

**Funding:** Evelyn Selby Stead Fund for Innovation, Vanderbilt University Medical Center

**Table of Contents:**

**Administrative Information**

**1.0 Roles and Responsibilities**

**2.0 Trial Sponsor**

**3.0 Role of study sponsor/funders**

**4.0 Composition of steering committee, data management teams**

**Study Schema**

1.0 **Background**
2.0 **Rationale and Specific Aims**

**2.1 Objectives**

3.0 **Animal Studies and Previous Human Studies**

**Methods**

4.0 **Inclusion/Exclusion Criteria and Setting**
5.0 **Enrollment/Randomization**
6.0 **Study Procedures**
7.0 **Risks of Investigational Agents/Devices (side effects)**
8.0 **Reporting of Adverse Events or Unanticipated Problems involving Risk to Participants or Others**
9.0 **Study Withdrawal/Discontinuation**
10.0 **Statistical Considerations**
11.0 **Privacy/Confidentiality Issues**
12.0 **Follow-up and Record Retention**
13.0 **Ethics and Dissemination**
14.0 **Appendices**

**Administrative Information**

**1.0 Roles and Responsibilities**

**Protocol contributors:** Study Principal Investigator, Colin Walsh, drafted, revised and received approval for the trial and its protocols. Dr. Walsh is affiliated with Vanderbilt University Medical Center, the host organization and study location. The Study Coordinator, Katelyn Robinson, assisted in organizing, delegating, and managing ongoing trial activities.

**2.0 Trial Sponsor:** None

**3.0 Role of study sponsor/funders:** Funders played no role in design and conduct of the study; collection, management, analysis, and interpretation of the data; preparation, review, or approval of the protocol or related manuscripts.

**4.0 Composition of steering committee, data management teams:** The trial team managed trial data in partnership with Health Information Technology (HealthIT) at the trial location. The ongoing review of study data by the trial team ensured that the clinical study could continue without jeopardizing participant safety and the continuing validity and scientific merit of the trial. Because the study team had developed, validated, and analyzed the precise Electronic Health Record (EHR) data needed to conduct the trial, the team led on data and safety monitoring. The PI and study coordinator were responsible for monitoring the safety of participants, and for ensuring that participants are not exposed to undue risk.

**Study Schema**

**1.0	Background**

Annually, 47,500 Americans die from suicide and over 800,000 die from suicide around the world. Suicide kills 132 Americans every day. The first step of suicide prevention is risk identification and prognostication prior to intervention. Traditional suicide risk prognostication relies on clinical judgment guided by validated instruments like the Columbia Suicide Severity Rating Scale (CSSRS), and the Ask Suicide-Screening Questions toolkit. Over the last few years, multiple validated statistical models have been published to improve suicide prognostication including work by Army STARRS, REACH VET, the Mental Health Research Network, our team at Vanderbilt University Medical Center (VUMC), and more.

Recent research suggests that statistical modeling combined with face-to-face screening might be better than either alone. However, the best risk prediction model possible must be made useful through tools like clinical decision support (CDS). CDS is an approach to engaging people with technology to improve the clinical care process. For example, a clinician might receive a prompt that will improve or adjust their clinical decision-making while prescribing a medication or ordering a test. Effective CDS would prompt a better decision than standard of care: the clinician making the decision unaided.

Prior literature outside suicide research has examined forms of CDS such as interruptive (e.g., alerts) and non-interruptive (e.g., static icons or visual cues) to inform contact isolation decisions, laboratory alerts, and blood transfusions. While that research has shown interruptive CDS tends to be more effective in changing behavior, this question has not been studied via Randomized Controlled Trial (RCT) in suicide prevention to our knowledge. Also, given significant concerns around false positives in suicide screening, demonstrating adequate performance of a non-interruptive CDS would support implementing a less burdensome and stigmatizing alert.

2.0 **Rationale and Specific Aims**

As above, the first step of suicide prevention is risk identification and prognostication. Researchers like our team have developed and validated predictive models that use routinely collected Electronic Health Record (EHR) data like past diagnoses and medications to predict future suicide attempt risk. Our model based in machine learning is known as the Vanderbilt Suicide Attempt and Ideation Likelihood (VSAIL). VSAIL has been validated prospectively and externally to predict suicide attempt risk with a number needed to screen (NNS) of 271 for suicide attempt and 23 for suicidal ideation. NNS is the number of people who need to receive a test result to prevent one outcome - lower NNS is better.

This study will evaluate the effectiveness of a CDS System called Vanderbilt Safecourse using VSAIL to prompt a novel Best Practice Alert (BPA) to prompt face-to-face screening with a validated suicide screening instrument like the Columbia Suicide Severity Rating Scale (CSSRS). We will compare the Interruptive BPA which includes the alert with a Storyboard icon that keeps the alert accessible if it is first "dismissed" with a Non-Interruptive Storyboard Only Prompt that does not interrupt clinicians but displays high risk predictions when present.

We seek to study if identifying patients at high predicted risk of suicide in clinical settings where suicide risk screening only happens sporadically, if at all, will include face-to-face screening rates and documentation of suicide risk assessment in their EHRs. With the Interruptive/Non-Interruptive design, we seek to test whether a non-interruptive alert is as effective in increasing face-to-face screening rates as an interruptive alert, which contributes more to "alert fatigue". If Non-Interruptive design is as effective as Interruptive, this advance would reduce potential burden of alerts in a clinical area in which risk identification might be stigmatized.

**2.1 Objectives**

We will measure two versions of the VSAIL-prompted CDS's effectiveness in real-world clinical settings to increase rates of face-to-face suicide risk screening. VSAIL requires only data already collected in routine clinical encounters and is calculated in real-time (seconds) at the start of a clinical visit (inpatient or outpatient) at VUMC.

*Primary Hypothesis*

We hypothesize the Interruptive CDS would be more effective at prompting in-person suicide risk assessment than Non-Interruptive (20% compared to 5%). Thus, we needed at least 75 patients in each arm to achieve 90% power with 5% probability of type I error.

3.0 **Animal Studies and Previous Human Studies**

No animal or human studies have been done.

Multiple analytic validation studies using deidentified data or clinical data repositories have been performed assessing the accuracy and validity of one study tool, the Vanderbilt Suicide Attempt and Ideation Likelihood (VSAIL).

1. Walsh, C.G., Johnson, K.B., Ripperger, M.A., Sperry, S., Harris, J., Clark, N., Fielstein, E., Novak, L., Robinson, K., Stead, W. "Prospective validation of a real-time predictive model of suicide attempt risk." JAMA Network Open. (2021)
2. Sulieman, L., Walsh, C.G. “Metrics for Predicting Suicide Attempts in Large, Imbalanced Clinical Biobank Data”. AMIA Joint Summits. (2020)
3. McKernan, L.C., Lenert, M., Crofford, L.J., Walsh, C.G. “Outpatient Engagement Lowers Predicted Risk of Suicide Attempts in Fibromyalgia.” Arthritis Care and Research. (2018)
4. Walsh, C.G., Ribeiro, J.D., Franklin J.C. “Predicting Suicide Attempts in Adolescents with Longitudinal Clinical Data and Machine Learning”. Journal of Child Psychology and Psychiatry. (2018)
5. Walsh, C.G., Ribeiro, J.D., Franklin J.C. “Predicting Risk of Suicide Attempts over Time through Machine Learning”. Clinical Psychological Science. (2017)
4.0 **Inclusion/Exclusion Criteria and Study Setting**

Patient Inclusion Criteria

1) > 18 years of age

2) A visit in Neurology (first phase) or other primary care setting at VUMC (second phase)

Patient Exclusion Criteria

1) A CSSRS conducted within one week of the visit in another care setting at VUMC

Provider Inclusion criteria for post-implementation interviews

1) Actively engaged with the BPA in regular patient care during the study period

Provider Exclusion criteria for post-implementation interviews

1) Providers who did not encounter the BPA

2) Providers no longer employed with VUMC

*Study Setting*

The study is conducted at Vanderbilt University Medical Center (VUMC), an academic medical center in the Mid-South of the United States. Within VUMC, the trial is conducted in ambulatory Neurology clinics, a non-behavioral health setting with increased suicide risk^40^ and variable suicide prevention workflows.

The study settings in this RCT include ambulatory Neurology clinics across three divisions: Neuro-Movement Disorders; Neuromuscular Disorders; Behavioral and Cognitive Neurology

5.0 **Enrollment/Randomization**

Randomization will be done in the Epic EHR, built-in functionality. Of patients in the highest predicted risk VSAIL tier (above a 2% predicted risk [Walsh et al, JAMA Open, 2021), one half will be randomized to the Interruptive Best Practice Alert (BPA) arm (intervention) and one half to the comparator arm - the Non-Interruptive Storyboard Only Prompt. For example, in outpatient Neurology, out of 60 patient visits per day, six patients on average fall into the highest risk tier and one half of those would be randomized to have the BPA pushed to the MD/NP provider providing care that day.

*Eligibility Criteria*

All patients proceeding for routine care in the Study Settings will be eligible for the RCT during its trial period.

6.0 **Study Procedures**

*Prediction and Randomization*

Prediction with VSAIL occurs in real-time at the start of a regularly scheduled, routine encounter in one of the participating study clinics. The patient’s registration for that visit prompts calculation of VSAIL which then populates a flowsheet in eStar. The VSAIL value above a predefined risk level (2% predicted risk based on our validation study, Walsh et al, JAMA Open, 2021 above) will prompt randomization within eStar to either the intervention arm or comparator arm. The randomization occurs within eStar, the Epic EHR version at the study site, through random number generation.

*Primary Outcome/Endpoint*

The primary outcome measured in this trial is Decision to Assess Suicide Risk, the clinician response to the intervention BPAs in each arm. This decision timepoint occurs when the practitioner interacts with the study CDS. In some instances, practitioners might "dismiss" alerts to act on them later. Each interaction including dismissals are recorded and analyzed to understand behavior around this study CDS.

*Secondary Outcomes/Endpoints*

Secondary trial outcomes include rates of thirty-day episodes of suicidal ideation, suicide attempt, rates of documented suicide risk assessment in clinical notes, psychiatric hospitalization, or emergency department utilization related to mental illness and/or suicide risk.

*Interventions*

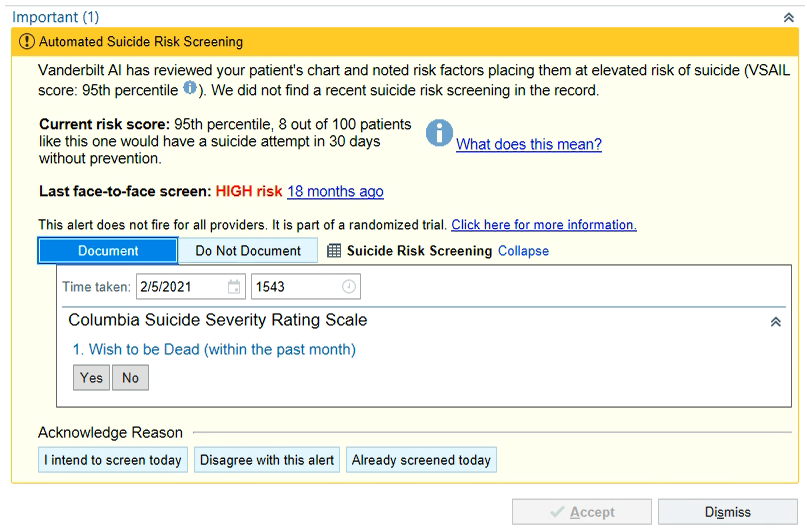
For patients randomized to the Interruptive VSAIL-prompted BPA, the Neurology MD clinician responsible for care in that encounter (determined in the usual course of care) will receive a BPA that shows the VSAIL score for that patient, e.g., 95^th^ percentile of risk which correlates with 3 out of 100 people like this one having a suicide attempt within 30 days. The BPA also shows the last face-to-face screening with the Columbia Suicide Severity Rating Scale (CSSRS), if one exists, with a link to that screening document itself. The BPA prompts an opportunity to “Document” or “Not Document” a new CSSRS instrument at that time if providers choose to do so. Finally, providers are able to share either i) Intent to Screen; ii) Disagreement with the BPA; or iii) that screening has already taken place. See Figure 1 for the visualization of this BPA.

Figure 1: VSAIL-prompted BPA, fake data used here (Interruptive Arm)

Dismissing the BPA (bottom right of Figure 1) leaves the Storyboard icon present in the eStar view, identical to the Non-Interruptive Arm described below. Hovering the mouse over the Storyboard icon reproduces the BPA alert and clicking on it permits the same functionality in the BPA as shown in Figure 1.

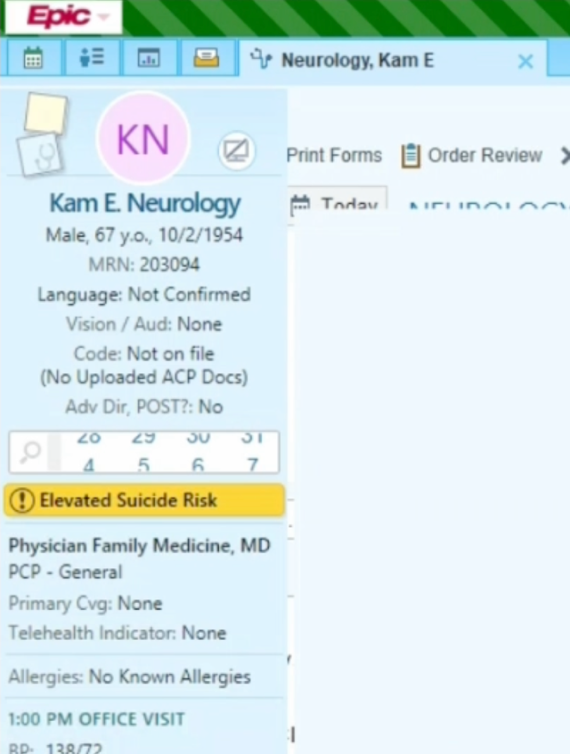
For patients randomized to the Non-Interruptive Storyboard Only Prompt (Non-Interruptive Arm), the provider will see an unobtrusive icon in the left side of the eStar window, the "Storyboard". The Storyboard is always present in eStar within an individual patient view. Hovering the mouse over the Storyboard icon reproduces the BPA alert and clicking on it permits the same functionality in the BPA as shown in Figure 1.

The VSAIL score will be available for all patients regardless of predicted risk tier in the study settings in the eStar flowsheet for equipoise but will not be pushed to providers outside the intervention arm. That is, providers who see value in the VSAIL risk score will be able to see it in eStar if they seek it out in the flowsheet.

Providers screening for or otherwise aware of VSAIL-predicted risk will have full discretion to treat suicidality clinically. The management of suicidality will follow standard of care at VUMC.

Figure 2: Storyboard icon (Passive Arm and "dismissed" BPA view)

Patients will experience routine care until providers screen for suicidality which they might not otherwise do without the VSAIL-prompted BPA for those identified at highest risk. That screening will take the form of the validated CSSRS or provider-chosen alternative (e.g., Patient Health Questionnaire-9 or direct questions about suicidal thinking posed by providers in their own way, the latter is consistent with known best practices to screen for suicidality in a direct and caring way; best practices do not dictate a single accepted screening instrument).

*Discontinuing Interventions*

Because the trial interventions occur at a single decision-making time point within a single study encounter, discontinuing interventions will not apply. The participating practitioners might dismiss/ignore the alerts or they might act on them. This decision is the primary study outcome, as above.

*Adherence to Intervention Protocols*

Adherence to the primary intervention outcome, decision to assess suicide risk, will be measured through direct interaction with the study decision support tools (see Interventions above). A secondary measure of adherence will be measuring whether suicide risk assessment is documented in clinical notes written as per routine clinical care in encounters in study settings during the trial period.

*Relevant concomitant care and interventions*

Decision to act on study CDS or not is left to practitioners in study settings. Any concomitant care is permitted at any point in the trial and during study encounters.

*Participant Timeline*

Randomization, interventions, and assessments all occur within each study encounter with no follow-up or additional interventions for either study participants (practitioners) or patients.

*Sample Size*

We hypothesize the Interruptive CDS would be more effective at prompting in-person suicide risk assessment than Non-Interruptive (20% compared to 5%). Sample size calculation indicates at least 75 patient encounters are needed in each arm to achieve 90% power with 5% probability of type I error.

*Recruitment*

Because of the pragmatic nature of this RCT, enrollment/randomization occurs at the time of check-in for routinely scheduled clinical care. No additional recruitment will be conducted. The rate of encounters is also stable as clinics tend to operate with consistent rates of clinical visits based on numbers of providers working in clinical settings.

*Post-Implementation Evaluation*

We will analyze the BPA utilization data routinely collected within the Epic EHR to identify providers who encountered the BPA in regular patient care during the study period. We will select a representative sample of encounters based on the BPA utilization data and contact providers through a general announcement to recruit up to 30 providers for follow-up semi-structured interviews. We will follow up as needed with the group of providers who meet inclusion criteria.

This study evaluates usage of a novel decision support system that does not exist in usual neurologic workflow. Providers might voice concerns or dislike of the system itself, issues related to suicide prevention practices in general in Neurology might be raised. No data specific to that provider's performance or practice will be collected that might increase risk of confidentiality. Safeguards to lessen this risk include deidentifying participants and redacting any identifying names inadvertently mentioned in interview sessions. Supervisors of participating employees will not be made aware of responses or decisions to participate. All data will be de-identified and all names will be changed to pseudonyms. All responses will be reported in aggregate.

Individual interviews will be conducted remotely via VUMC-approved secure HIPAA compliant video conferencing platform (e.g., Teams), audio/video recorded and transcribed, and last 30-60 minutes. Recordings will be stored on VUMC-approved secure servers (VUMC OneDrive or Sharepoint) and only authorized study personnel will have access.

Participants who complete an interview will be compensated with a $50 electronic Amazon gift card which they will receive via email no later than one week after the interview.

7.0 **Risks**

This protocol presents minimal risks to participants. Adverse events are not anticipated. Risks include discomfort among both providers and patients in discussing the sensitive but important topic of suicide. The risk model is not perfect and might not identify risk in those who are at risk – it is not intended to preempt providers screening any patient (or all patients) for suicidality if they choose to do so. The risk model might also identify risk in those not at risk, leading to questions of suicide screening being asked unnecessarily.

8.0 **Reporting of Adverse Events or Unanticipated Problems involving Risk to Participants or Others**

*Harms*

The PI is responsible for monitoring study data, assuring protocol compliance, and conducting safety reviews. Because of the pragmatic nature of the study interventions (CDS decision aids), adverse events or other problems are not anticipated. In the unlikely event that such events occur, unanticipated problems involving risks to subjects or others that are i) serious, ii) unanticipated, and iii) possibly or definitely related to research procedures will be reported immediately to the PI, and in writing within 7 days to the VUMC IRB. The PI will apprise fellow investigators and study personnel of such adverse events that occur during the conduct of this research project through regular study meetings. Continuing review reports will be submitted annually to the IRB summarizing study progress, adverse events, complaints about the research or withdrawals, and any protocol violations.

We will follow the VUMC Human Research Protections Program (VHRPP) guidelines that require investigators to promptly notify the IRB within 7 days of the occurrence when such unexpected Adverse Events (AEs) occur. These are events that could possibly be related to the intervention, or occur more frequently or are more severe than anticipated. VHRPP requires that any AE that is unexpected and related or possibly related to the research be reported. Adverse events not meeting this definition will be reported at the time of continuing review.

Unanticipated problems involving risks to subjects or others will be reported immediately, within twenty-four (24) hours of their discovery to the PI, and in writing within 7 days to the IRB. The PI will apprise fellow investigators and study personnel of all AEs that occur during the conduct of this research project through regular study meetings. Annual reports will be submitted by the study coordinator to the responsible IRBs summarizing study progress, AEs, complaints about the research, and any protocol violations.

*Auditing*

Study data will be analyzed by the PI and study team every three months for 1) continued validity of VSAIL to predict suicide attempt; 2) proper functioning of the VSAIL model [e.g., ensuring all predictors continue to be calculated appropriately for patients]; 3) rates of suicidal ideation and suicide attempt in patients randomized in the study. Because this study necessitates close collaboration with providers receiving BPAs, the study team will have monthly small-group meetings with providers in study sites at VUMC to assess for problems or perceived AEs related to the study protocol, though none are anticipated.

9.0 **Study Withdrawal/Discontinuation**

Providers may dismiss the BPA and not act on its recommendations at any time. No other intervention is planned. Patients would be unaware of the providers’ decisions in that case unless the provider chooses independently to discuss the BPA with them.

10.0 **Statistical Considerations**

In consultation with the study biostatistican (Dr. Cindy Chen) and Learning Health System (Dr. Christopher Lindsell), the study team determined the following power analysis.

On average at a predicted risk threshold of 2% predicted risk (the highest risk tier in settings without universal screening), 6 patients per day are anticipated in the highest risk tier. Randomizing half to the intervention arm and half to the comparator arm and 15 patients a week per arm, we anticipate 20% of the BPA prompts will be associated with face-to-face affirmation compared to 5% in the comparator arm, which means we need 75 patients in each arm to achieve 90% power with 5% type I error, 200 total, which will require fewer than six months to accumulate. Because prior suicide attempt is rare in this cohort based on diagnostic code ascertainment, simple randomization as above is justified.

11.0 **Privacy/Confidentiality Issues**

All study experiments occur solely in the context of already-scheduled clinical care and among patients and providers already intending to interact for purposes of healthcare delivery. No data outside that patient’s health record are used in the calculation of VSAIL or in study conduct. Because these interactions occur solely in routine healthcare encounters, the same confidentiality rules apply as in standard of care.

Study data are secured on servers in the VUMC data center or in VUMC Health Information Technology production systems only. These systems are secured via personal, physical, and technical controls including their invisibility to networks outside the VUMC firewall. User credentials are maintained by the VUMC Active Directory and Information Technology and are necessary to access any study data. Only those credentialed and on an approved IRB will have permissions to access study data on these servers. No paper records will be generated nor recorded in this study. All data collection will be digital and will leverage operational clinical systems and EHRs in their generation. No new data collection mechanisms will be required and therefore no new vulnerabilities generated.

12.0 **Follow-up and Record Retention**

The study will last six months.

As per IRB policy VI.B, all study data will be maintained for six (6) years from the date of the last use of study Protected Health Information (PHI) - at the conclusion of the trial period to identify patients in need of outcome data collection (which is based in EHR queries only).

13.0 **Ethics and Dissemination**

*Institutional Review Board/Ethics Review Board Approval*

This study was approved by the VUMC IRB #210865.

*Protocol Amendments*

Changes to trial protocols including eligibility criteria, outcomes, or analyses will be communicated to the governing IRB, updated in national registration (ClinicalTrials.gov), clinical trial site leadership and participating clinicians within one calendar day.

*Waiver of Consent – in-clinic preventive CDS*

Waiver of consent was approved by the VUMC IRB (#210865) for the in-clinic CDS components of the trial. This study aims to measure the effectiveness of a risk model-driven BPA on provider behavior. Multiple factors contribute to impracticability to obtain consent. Consenting only those identified at highest risk would itself introduce selection bias and might alter behavior of staff/providers aware that consent had taken place in a manner that would alter study results. Additionally, the timing of discussing suicidality should be and will be left to providers at the appropriate time in a clinical visit, ideally after some rapport had been established. Informed consent at the start of the visit disrupts that intended workflow. Finally, in the most severe cases of suicidality, decisional capacity might be affected making patient informed consent impossible. Waiver prevents that issue from i) disrupting care for those patients and ii) unduly affecting the study itself.

*Waiver of Consent – minimal risk*

This study does not involve an investigational intervention given or administered directly to patients. This study poses no greater than minimal risk to study participants. VUMC has become a leader in pragmatic, minimal risk clinical trial designs like this one and we have engaged the Learning Health System for high-level guidance in study design. Our study team benefits greatly from the experiences and expertise of our colleagues who have completed similar studies under waiver of informed consent. The potential benefit of this study would come through better provision of screening to those at risk who would not otherwise be screened in current state. Waiver of consent will have no impact on privacy for patients or providers because the CDS will be applied only in routine encounters already scheduled for unrelated reasons, and no data from this study exist outside the bounds of the individuals' EHRs.

*Informed Consent – Post-trial qualitative evaluation*

For those providers who choose to participate, we will email each participant the link to RedCap eConsent form in advance of the interview. The RedCap eConsent framework allows participants the ability to download a copy of their signed consent form. At the start of the interview session, we will review the elements of the informed consent and give participants an opportunity to ask questions of the researcher. During this consent conversation, providers will be reminded that participation is optional and that they may choose to withdraw at any time without any consequences.

*Confidentiality*

Data will be collected from the EHR and stored securely in the VUMC Data Center on servers owned by the study team for later analysis. These systems are designed to securely store Protected Health Information and include encrypted storage, multifactor authentication, and both physical (locked doors, secure facility) and technical (active directory user authentication) controls. Protected Health Information, while necessary to conduct study analyses, will never be exported from these servers or shared.

Data to be collected include the VSAIL-produced probability, the predictors for that probability including historical counts of diagnoses, demographics (age, sex, race, zip code), medication counts, visit utilization (counts of outpatient,
inpatient and emergency department visits) for the preceding five years. We will also collect documented and coded suicidality including suicidal ideation (a single ICD10 code) and suicide attempt (a reference set of ICD10 codes used by our team and others from the CDC Center for National Health Statistics). We will also collect decision support usage statistics included face-to-face screening rates (the Columbia SSRS), and responses to the BPA: 1) "intent to screen" based on the alert; 2) "already screened for suicidality in this visit"; 3) "I disagree with this

alert". Additional data will be collected from the same EHR data to evaluate secondary outcome and process metrics following the randomization: emergency department utilization; psychiatric consultation; documentation of suicide
screening in clinical notes.

For provider interviews post-trial, we will deidentify all interview data and store data in licensed storage with companies with which VUMC has a Business Associate Agreement (BAA), e.g., Microsoft OneDrive. Data will include impressions of the CDS, its role in decision-making, barriers to its use, and perceived strengths and problems in its design.

*Declaration of Interests*

No conflicts of interest exist, financial or otherwise, for any members of the study team relevant to this research.

*Access to data*

Only two study team members have access to raw EHR data needed to conduct study analyses: Colin Walsh (PI), Michael Ripperger (developer/analyst).

Only members of HealthIT with Physician Builder credentials necessary to design and implement the trial CDS might access EHR data within our vendor EHR: Ashley Spann (Physician Builder), Dan Albert (HealthIT).

*Ancillary and Post-trial care*

Not applicable to this study.

*Dissemination Policy*

Our team will disseminate trial findings through peer-reviewed journals and national conferences in biomedical informatics, psychology, psychiatry, neurology, and potentially emergency medicine and internal medicine. Where possible, the team will preprint our manuscripts at the start of peer review for more transparent dissemination of our results. We will pursue open access publication where available to maximize dissemination.

No professional writers will be employed in dissemination.

Because of the sensitive nature of the PHI needed to conduct this trial, public access to participant-level data will not be possible. Statistical code and the full protocol for the study (this document) will be disseminated with peer-reviewed publication as appropriate.

**14.0 Appendices**

*Informed consent materials*

Waiver of consent, as above

*Biological Specimens*

Not applicable
