## Supplementary material for "Randomized Controlled Comparative Effectiveness Trial of Risk Model-Guided Clinical Decision Support for Suicide Screening": Study Protocol

eSupplement, Interruptive and Non-Interruptive CDS User Interfaces

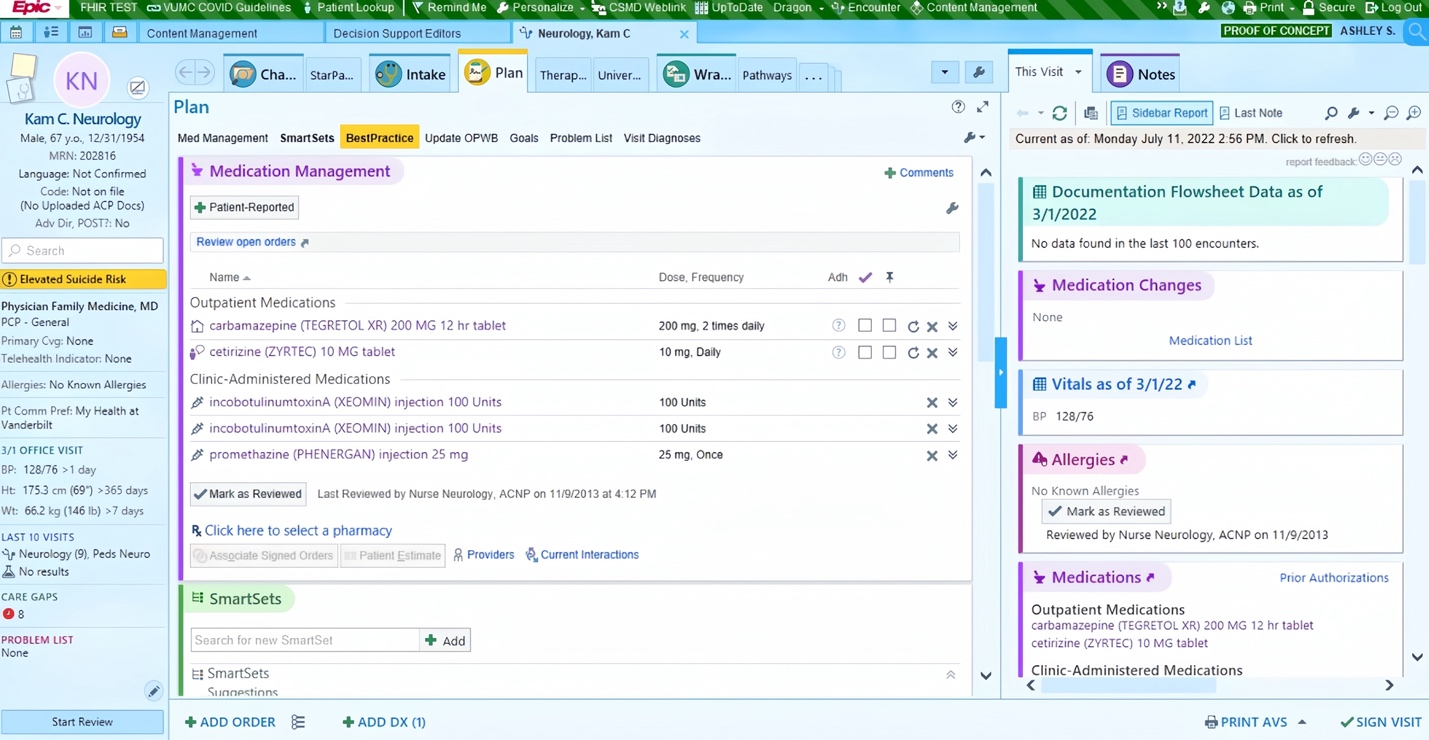

Figure 1a: Non-Interruptive CDS, “Elevated Suicide Risk” icon shown in patient panel

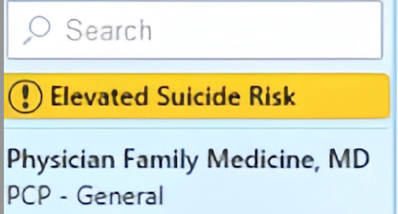

Figure 1b: Non-Interruptive CDS/Storyboard icon shown, detail

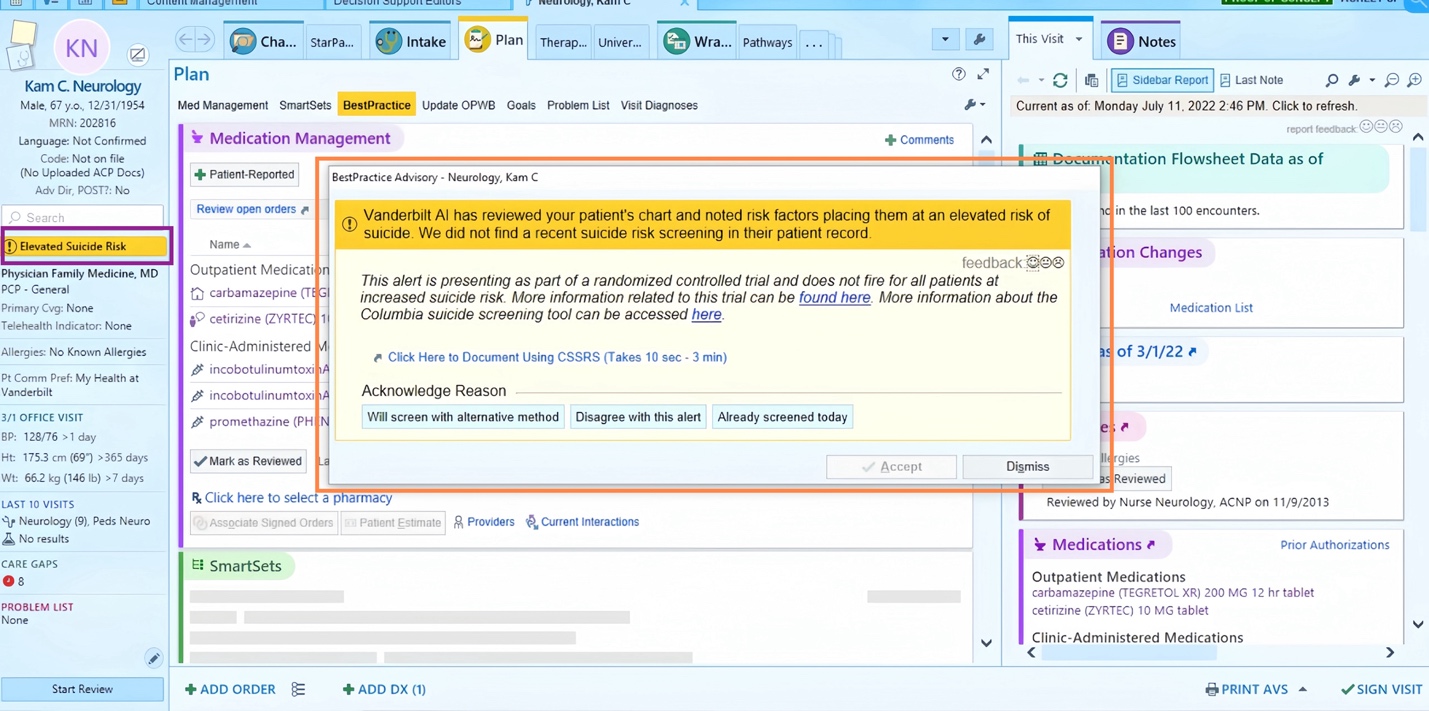

Figure 2a: Interruptive CDS, BPA highlighted (orange) and patient panel icon highlighted (purple). Fake patient data shown.

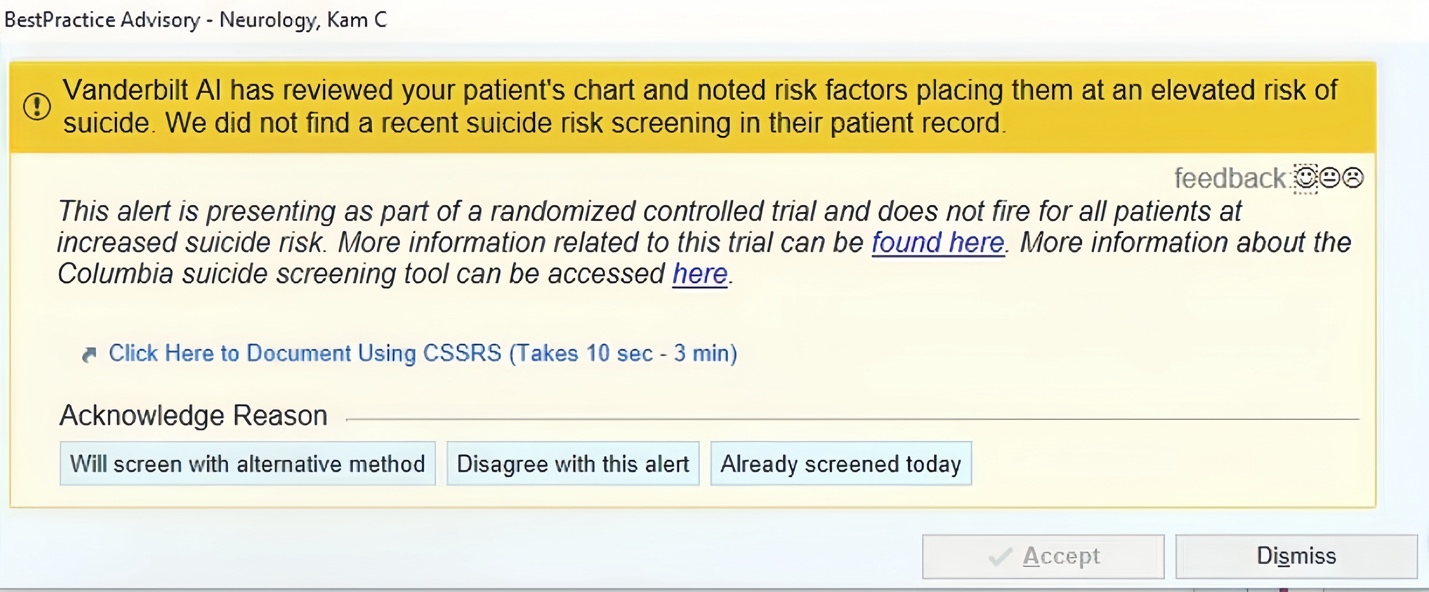

Figure 2b: Interruptive CDS BPA shown, detail

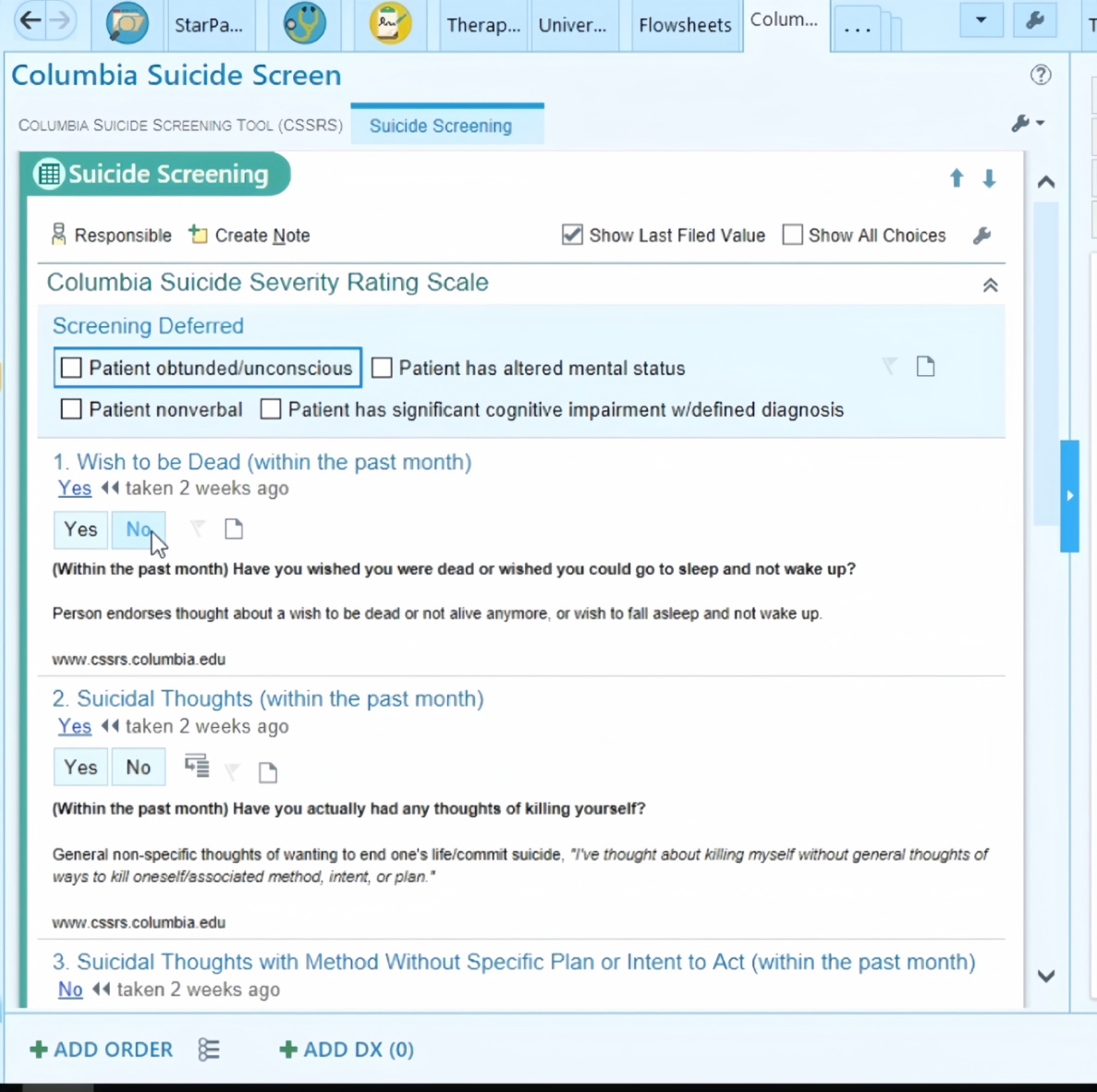

Figure 3: The CSSRS Custom Form (excerpt) developed for this RCT

**Methods, seed terms derived from RCT chart review**

Seed terms extracted in RCT chart review with wildcards denoted as %: “suicid%”; “ SI.” “ SI “ “SI/HI” “phq” “cssrs”.

**Comments input to CDS by Theme**

| Theme | Comment |
| --- | --- |
| Screened (N=32) | administered PHQ-9, item #9 was 0; clinical interview prompted for SI/SA, denied by pt and informant |
|  | BDI-II = 13 (minimal) |
|  | C-SSRS administered - hard copy will be uploaded to EMR |
|  | C-SSRS administered because of this alert |
|  | C-SSRS performed because of this alert - form will be uploaded to EMR |
|  | C-SSRS performed following alert trigger |
|  | C-SSRS screen negative on day of encounter |
|  | Denied current or past SI or suicidal behavior |
|  | denied suicidal ideation on Beck Depression Inventory as well as on interview. |
|  | Denies SI [two identical comments] |
|  | gds15 |
|  | Interview |
|  | Negative [two identical comments] |
|  | No SI |
|  | no SI at present |
|  | no SI/HI |
|  | Not suicidal |
|  | phq9 [three identical comments] |
|  | Reports no SI |
|  | Screen negative |
|  | Screened |
|  | screened because of this alert |
|  | screened day of visit |
|  | Screened during appointment on <DATE>. Will be documented in neuropsych report |
|  | Screened during clinical interview, No SI indiciated [sic]. |
|  | screened in part because of this alert |
|  | screening initiated because of this alert |
|  | will screen with C-SSRS, given this alert |
| Inappropriate for patient (N=4) | no risk factors [two identical comments] |
|  | <Patient is non-verbal and "unable to express suicidality"... "I am uncertain of how to appropriately screen for suicidality"> |
|  | Patient with severe dementia |
|  | inaccurate |
| Deferral (N=2) | I am opening the chart for the first time. I don't know anything about the pt |
|  | patient seen with resident |
| Context (N=1) | did this the date of pt encounter; opening chart for addendum |

*Table: Physician comments to BPAs, [sic] noted and any study team redaction noted in <>*
